# Precision of Mammogram and USG based evaluation of NAST response in breast cancer and its impact on oncoplastic surgery decisions

**DOI:** 10.1101/2021.08.06.21261694

**Authors:** Pooja Deshpande, Santosh Dixit, Devaki Kelkar, Nutan Gangurde, Shahin Shaikh, Sanket Nagarkar, Smeeta Nare, Laleh Busheri, Chaitanyanand Koppiker, Beenu Varghese

**Author notes:** **Corresponding author:** Dr Beenu Verghese and Dr Chaitanyanand Koppiker. This manuscript is dedicated to the memory of Dr Pooja Deshpande who was involved in the conceptualization, design and development of this research project.

## Abstract

**Background:** Precise prediction of residual tumour size following neoadjuvant systemic therapy for breast cancer is crucial in assessing response and surgical decision making. Our study is aimed at assessing the performance of conventional imaging modalities like ultrasound and mammography in predicting the residual tumour size after neoadjuvant systemic therapy and in evaluating the impact of imaging on the surgical outcomes.

**Methods:** We retrospectively compared the tumour size measured by ultrasonography and mammography and the residual tumour size on final histopathology in 109 patients. Concordance was defined as a size difference within 25% of the histopathology size. We also looked at the distribution of concordance between different T status and molecular subtypes, accuracy of USG in predicting pathological complete response and axillary lymph nodal metastasis and also surgical outcomes in the discordant cases.

**Results:** The concordance rates of mammography and ultrasonography were 68.2% and 52.3% respectively without statistically significant difference between the two modalities (p = 0.081). Combination of both the modalities had a concordance rate of 57.8%. Ultrasonography had accuracy of 81.7% for predicting pathological complete response and 79.8% for predicting axillary nodal metastasis. We did not identify any influence of histologic subtype on the associations between preoperative measurements and pathology size or the accuracy for detecting pathological complete response (p values 0.43 and 0.46 respectively). In 12 cases, the radiology-pathology discordance could have led to large excision volume surgeries. In the overall cohort, recurrence free survival and overall survival rates at median follow up of 19.1 month were 87.2% and 95.4% respectively.

**Conclusions:** Ultrasound and mammography showed moderate concordance with pathology for estimation of the residual tumour size without any significant difference in the performance between the two. Despite the moderate concordance, surgical outcomes were fairly well managed in the discordant cases with the oncoplastic surgical techniques. Our study highlights the usefulness of the cheaper and widely available conventional imaging modalities in the developing countries where the cost of treatment is to be contained.

## INTRODUCTION

neoadjuvant systemic therapy (NAST) is increasingly being used in Early Stage Breast Cancer (EBC), besides its use in advanced disease. NAST confers several advantages, a major one being reduction of tumour burden. This can convert potential mastectomy cases into breast conservation surgery (BCS) or reducing the extent of excision in EBC that allows the surgeon to perform the more cosmetically desirable oncoplastic breast surgeries (OBS) (Bhattacharyya *et al*. 2008). Furthermore, attainment of post-NAST pathological complete response (pCR) of the primary tumour, is associated with treatment response in clinically positive axillary nodes. There is increasing evidence that in cases with a clinically positive node that shows complete response on post-NAST imaging, one can avoid complete axillary lymph nodal dissection (ALND) and instead do a sentinel lymph node biopsy (SLNB) or a targeted axillary dissection (TAD). These collectively contribute to the prognostic significance and is associated with improved disease-free survival post-NAST (Fowler *et al*. 2017).

Predicting residual tumor size after NAST is especially challenging due to post-therapy reactive inflammation, fibrosis, necrosis and fragmentation. However, accurate prediction of residual tumor size is essential for surgical decision making and to assess response to treatment (Keune *et al*. 2010). The main goals of the surgeon in doing BCS are to obtain tumour free margins, this in turn reduces local recurrence rates and enables attainment of cosmetically desirable results. The latter can be partly achieved by removing the least amount of normal tissue as possible. With the adaptation of the OBS techniques, it is now possible to excise larger volumes of breast tissue and still achieve good cosmesis (Clough *et al*. 2003). An additional challenge in the post-NAST scenario is accurate identification of location of the residual tumour, especially in cases where there have been good responses to NAST (Warren *et al*. 2004a; Yeh *et al*. 2005; Keune *et al*. 2010).

Residual tumor size after NAST is assessed by clinical breast examination (CBE) and with imaging modalities like mammography (MG), ultrasonography (USG) and magnetic resonance imaging (MRI). A meta-analysis by Marinovich et al (Marinovich *et al*. 2013) concluded that MRI had high sensitivity of 92% but rather low specificity of 60% to correctly detect residual tumor after NAST. Additionally, individual patient data (IPD) meta-analysis by Marinovich et al (Marinovich *et al*. 2015) concluded that there is an overall better agreement between MRI and pathology, suggesting that MRI is the more appropriate assessment method. However, it is possible that a combination of USG and clinical examination may be superior to either test being used individually.

Our study was aimed at determining the accuracy of conventional modalities which are widely available and cost effective like USG and MG in assessing the residual tumor size after NAST. We examined the impact on surgical decisions based on the residual tumor size provided by conventional imaging.

## MATERIALS AND METHODS

### Study Population/Cohort

We did a retrospective study on 109 patients who received NAST in the form of neoadjuvant chemotherapy (NACT) or hormonal therapy (NAHT) for invasive breast cancer, and underwent subsequent definite surgery during the period of January 2014 to May 2019. The study was approved by the local Ethics Committee and by the host institution’s (Orchids Breast Health Clinic and Prashanti Cancer Care Mission) Research and Development office. All participants provided written informed consent.

The patients who did not complete NAST or who underwent definitive surgery without undergoing NAST and patients with stage IV disease were excluded from the study. The patients in whom neither preoperative MG nor USG images were available (imaging done elsewhere or no records available) were also excluded.

### Radiology Imaging Mammography

Mammography was performed on the Siemens Inspiron digital breast tomosynthesis (DBT) unit. 2D and DBT Images were acquired in craniocaudal (CC) and mediolateral oblique (MLO) views. Additional views like spot magnification, exaggerated CC views were also acquired if needed.

### Ultrasonography

The USG examination was performed on the Siemens acuson S 2000 with a 9 -14 Mhz frequency linear transducer. All USG examinations were performed by one of the two breast radiologists who had between 6 and 21 years of breast USG experience. Besides grey scale imaging, colour doppler and elastography were also used in the evaluation of the lesions.

### Imaging Protocol

The imaging protocol included a baseline (pre-NAST) MG followed by an USG of the breast. Pre-NAST mammograms were available in 70 cases and pre-NAST USG images were available in 103 cases. In the rest of the cases, pre-NAST imaging was done elsewhere and was not repeated at our clinic due to cost issues. The ipsilateral axilla was examined in all cases to look for Level I & II axillary nodes. If abnormal axillary nodes were identified at these levels, evaluation of level III nodes was done.

Follow up interval USG for the primary breast lesion and abnormal axillary nodes if any, was scheduled following every two cycles of NAST. The scheduling of the interval examination was also determined by the clinical examination. An interval MG was only obtained if there was difficulty in assessing the lesion on USG.

Placement of clips into the tumour bed was done at an appropriate time point on follow up imaging during NAST, depending on the tumour response. Clip placement into the nodes was done at the time of FNAB in a few cases.(Case 2)

Preoperative MG and USG examinations were performed within 14 days prior to the surgery date. Preoperative MG images were available in only 70 cases as only analogue mammography system was available in our clinic during the year 2014-2015 and hence images were not available on the PACS (Picture Archiving and Communication System). Besides we were not performing preoperative mammography in all cases during the first 3 years of the study. Preoperative USG was available in all the 109 cases.

### Radiological Analysis

MG and USG Image interpretation was done using the fifth edition of the Breast Imaging Reporting and Data System (BI-RADS) lexicon. The longest unidimensional preoperative tumor size measurements (USG and or MG) before and after NAST were recorded. In cases with satellite lesions the total extent of the abnormality was recorded. In case of multifocal/multicentric tumors, the longest dimension of the largest tumor was documented. The maximum cortical thickness of the axillary lymph nodes was documented. If lymph nodes showed complete loss of fatty hila, node measurements were taken in two orthogonal planes. Also, the number of abnormal axillary nodes and evidence of peri nodal infiltration were documented. In 4 patients in whom visualization of tumor margins was difficult on 2D mammogram and DBT, only the USG dimensions were taken into consideration.

### Clinicopathological Analysis

The clinicopathological data of these patients including patient demographics, treatment details, baseline as well as surgical HPE were collected through review of the surgical and pathological records.

### Baseline Histopathology

All patients underwent a presurgical diagnosis of tumor histology by core needle biopsy (CNB) with a 14G core biopsy needle at the time of initial imaging. Abnormal nodes were also biopsied either with a core biopsy or a FNAC. The histological type and grade of the tumour were documented. Immunohistochemistry (IHC) reports of Estrogen receptor (ER), Progesterone receptor (PR) and Human epidermal growth factor receptor 2 (HER2) status were available for these tumours. The Fluorescent in situ Hybridization (FISH) test was done for cases with equivocal HER2 (2+) reports. Molecular subtypes were documented as hormone receptor (HR) positive (ER/PR positive, HER2 +/-), HER2 enriched (ER and PR negative, HER2 positive) and triple negative breast cancer (TNBC) (ER, PR and HER2 negative).

### Surgical Histopathology

Pathologic tumor size was confirmed histologically on formalin-fixed tissue from serial sections made along the main axis of the tumor. Longest dimension of the invasive tumor was noted. In cases with mixed invasive ductal carcinoma (IDC) and DCIS, both components were documented separately. In cases where scattered foci of residual disease were found, the total extent of the scattered foci was estimated. pCR was defined as the total absence of invasive cancer in the breast as well as lymph nodes (ypT0/isN0), in situ disease was not considered while reporting pCR (Hortobagyi *et al*. 2017).

### Statistical Analysis

Cohort data was analyzed in R 4.0 on a Microsoft Windows 10 platform. Measurements of longest tumour dimension were considered as concordant when the percentage ratio of the difference in measurements by the imaging modality (USG/MG and MG+USG) and histopathology was less than or equal to 25% and discordant when greater than 25%. Discordant cases were further subclassified as overestimation when imaging measurement was larger than 25% and underestimation when it was smaller than 25% histopathology size. Mammogram and Ultrasound data were combined such that the longest available length from either modality was taken (hence, MG + USG). This combined column was analyzed as Imaging data. The resulting distributions were tested for significance over different features of the residual disease using ChiSquare Test (Table 2). Sensitivity analysis was done using the caret package in R (Kuhn 2020).

Positive events were taken as detection of residual disease and a negative event was resolution of disease *i*.*e*., PCR, N0, T0. Thus,

⍰ PPV: the ability to correctly predict the presence of residual disease on final HPE.
⍰ NPV: the ability to correctly predict resolution of disease on final HPE pCR.
⍰ Sensitivity: percentage of correctly predicted residual disease from all actual residual disease cases
⍰ Specificity: percentage correctly predicted resolution of disease from all cases of resolved disease.

Mathematically, the evaluation metrics were calculated in caret as follows:

⍰ PPV = TP/ (TP + FP)
⍰ NPV= TN/ (TN + FN)
⍰ Sensitivity = TP/(TP+FN)
⍰ Specificity = TN/(TN+FP)

Finally, Accuracy was calculated as the sum of true positives and negatives to the total number of samples measured. Cohens unweighted kappa is calculated as the ratio of difference between probabilities of observed and expected positives to the highest probability of expected positives. 95% confidence limits were calculated as a binomial proportion confidence interval as described by Kwiecien et al. Bland-Altman analysis was carried out using the blandr package (Datta 2017). 95% confidence limits were used. For each case, the longest dimension of the tumour obtained in the imaging modality was compared to the corresponding histopathology longest dimension. A p value of 0.05 or less was considered statistically significant.

## Results

A total of 109 patients with histologically proven breast cancer who underwent NAST followed by definitive surgery were retrospectively evaluated. In our study, the age group of patients ranged from 23 to 78 years with a mean age of 50.9 ± 11.3 years. Histology of the tumours were IDC in 100 (91.7%), ILC in 4 (3.7%) and IDC with DCIS in 5 (4.6%) cases. Analysis of the demographic data of the cohort is depicted in **Table 1**.

**Table 1:**
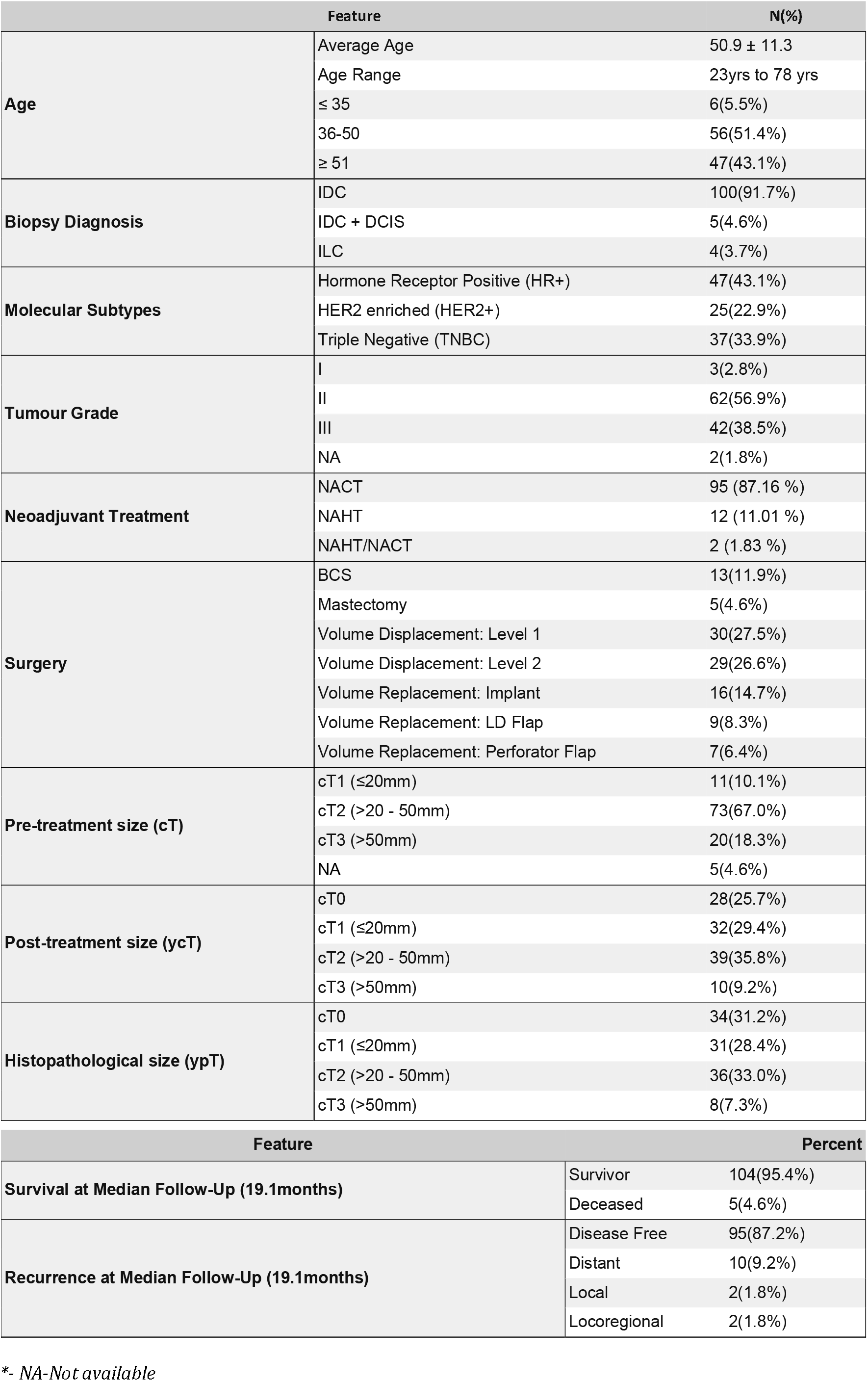
Demography of Cohort (n=109)

The tumour measurements by MG were not possible in 4 out of 70 cases due to extremely dense breasts. The tumour sizes measured by MG were concordant with the final HPE sizes in 45 out of 66 (68.2%) cases, underestimation was noted in 11(16.7%) cases while overestimation was noted in 10 (15.2%) cases. On the other hand, out of the 109 patients with USG, 57 (52.3%) cases were concordant, 21 (19.3%) cases were underestimated and 31 (28.4%) cases were overestimated. However, the difference between the performances of the two modalities was not statistically significant (p value-0.081). Combination of both the modalities had a concordance rate of 57.8% **(Table 2 and Figure 2a)**. Comparisons of MG and USG in the same patients showed similar concordance rates 68.2% and 53% respectively, without statistically significant difference between the two (p value-0.2).

**Table 2:**
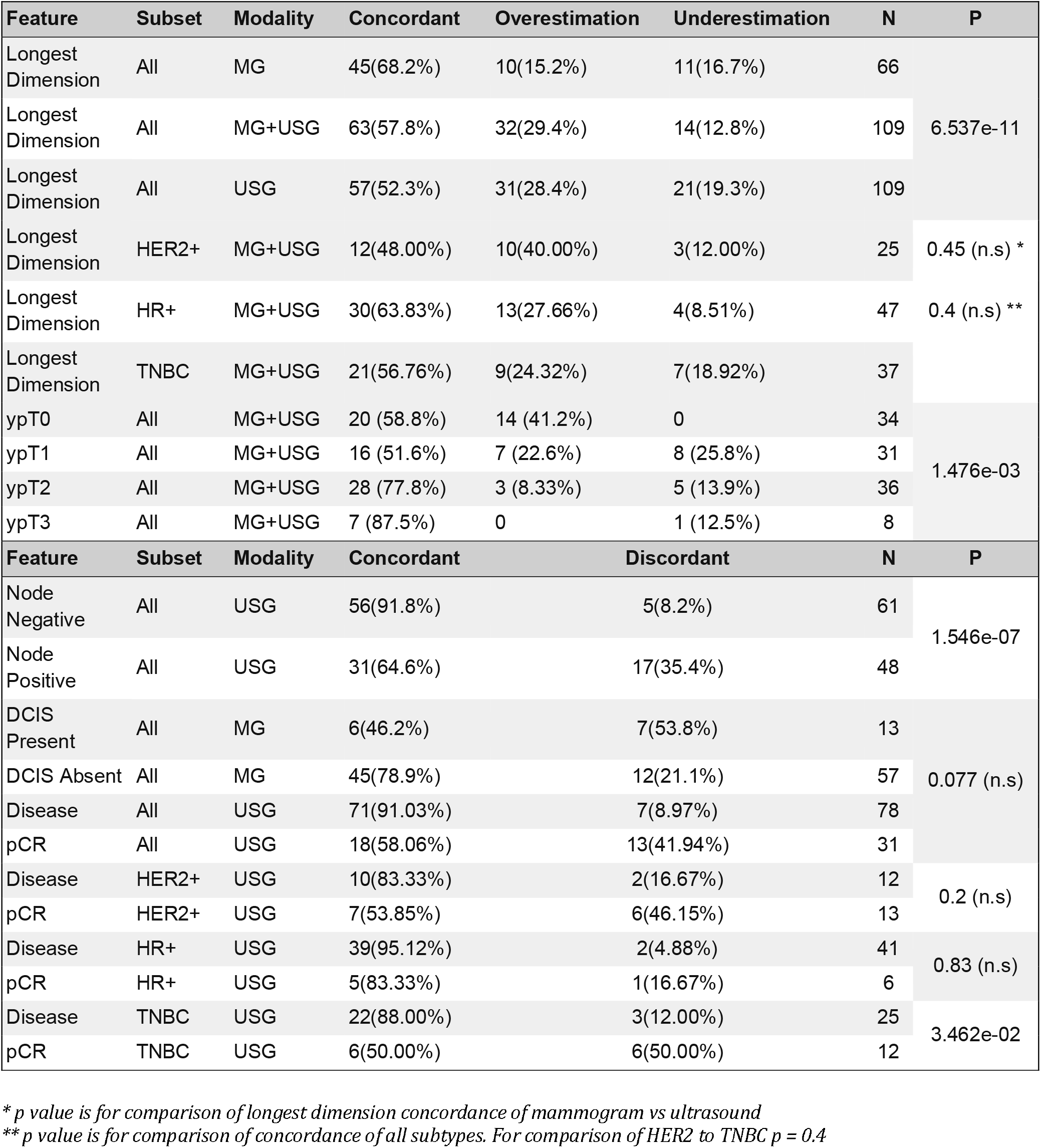
Concordance and Discordance of Imaging Modalities with Various Features of Residual Disease.

**Figure 1.**
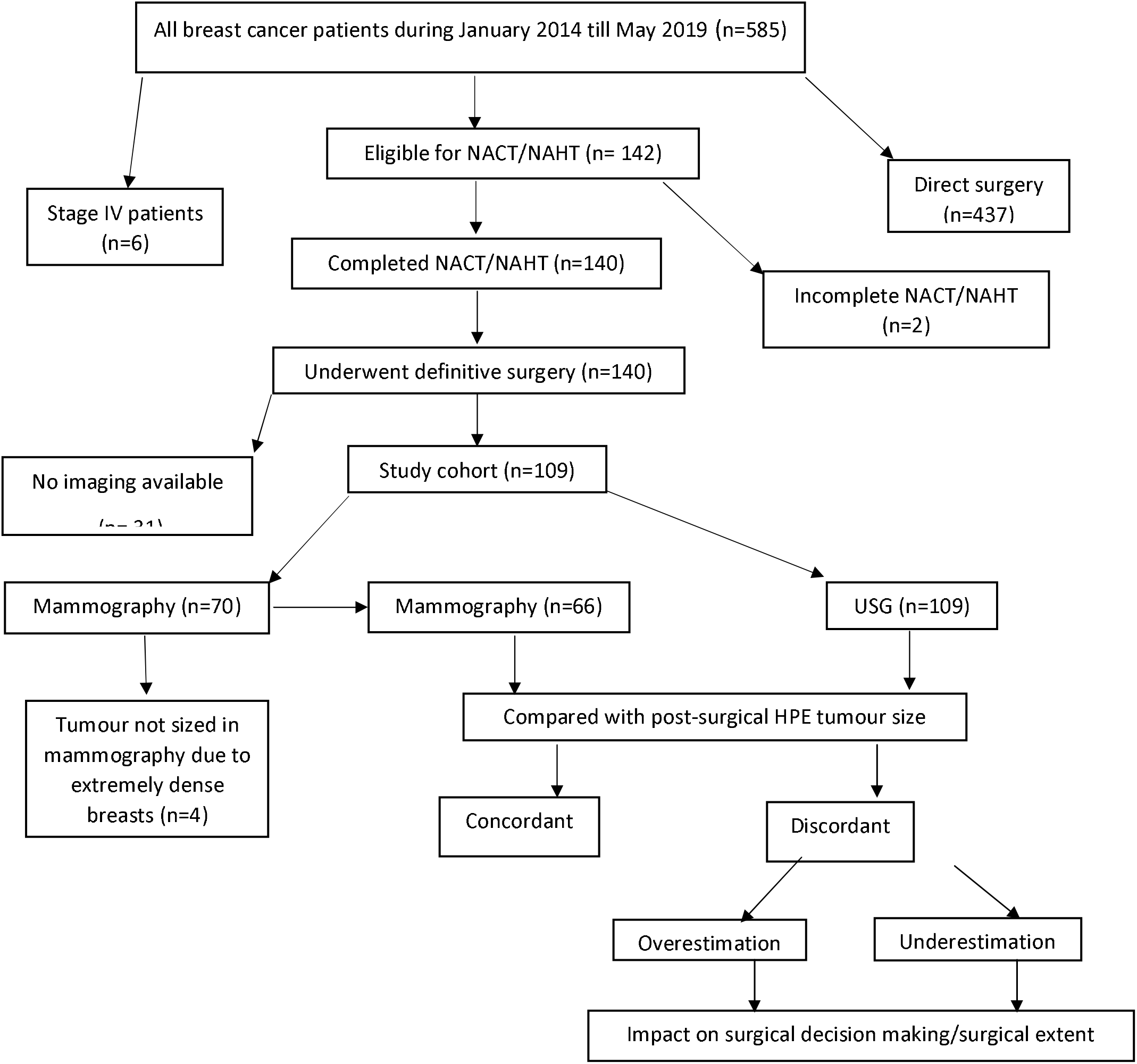

**Figure 2:**
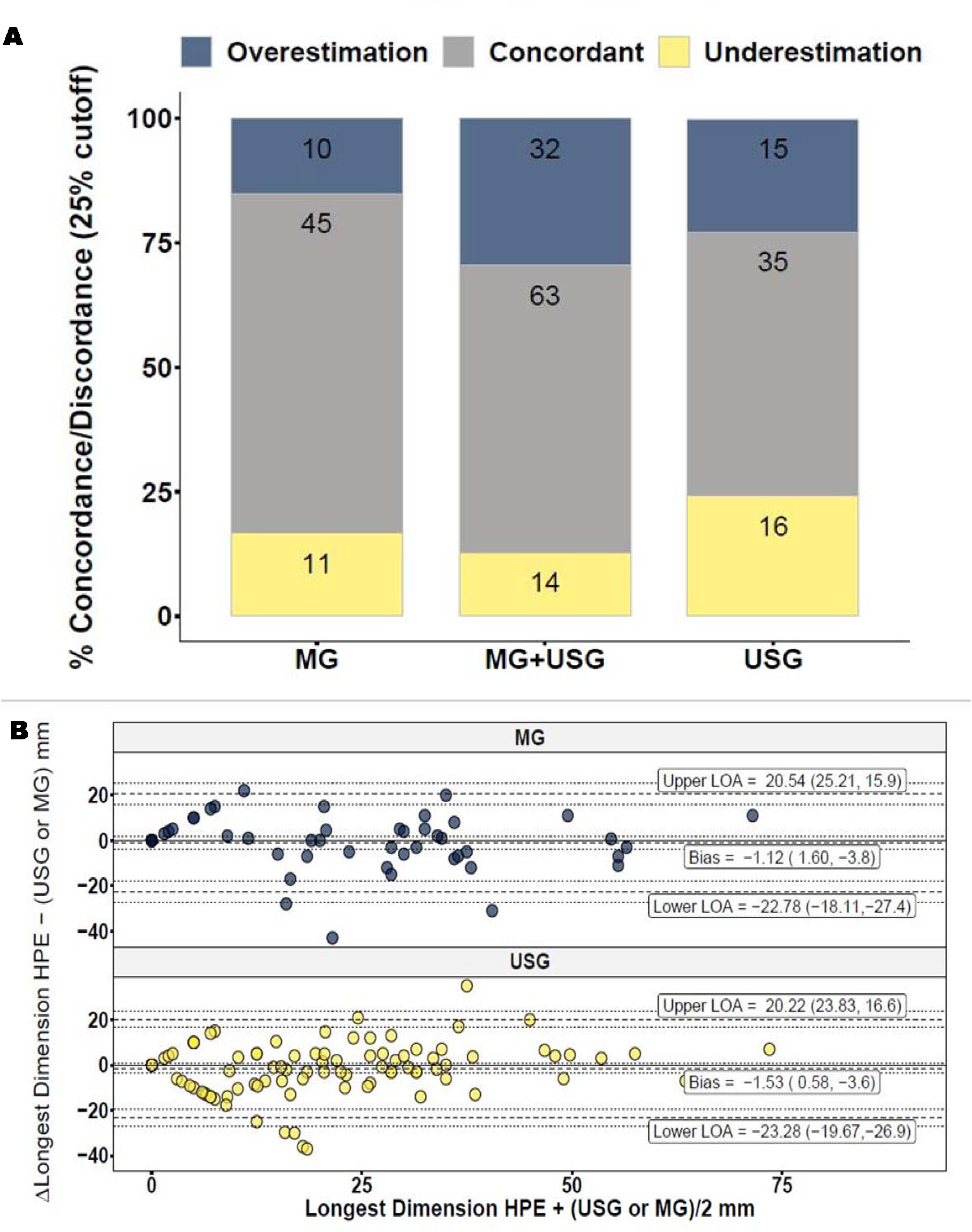

Bland-Altman analysisof both modalities. (**Figure 2b and Table 3)** showed a negative bias, -1.53 (0.58,-3.64) mm and -1.12 (1.6,-3.83) mm for USG and MG respectively. The combined imaging modality (MG+USG) has an increased negative bias of -3.52 (−1.45,-5.59). Paired two-sided t-test did not show any significant difference in the mean differences of these modalities from HPE (p = 0.22). The two modalities therefore are not significantly different across the range of tumour dimensions being studied. The spread of the Bland-Altman limits of agreement (Upper LOA – Lower LOA) varied over a small range from a maximum of 43.8 mm for MG and a minimum of 42.8 mm for combined MG + USG. Visual inspection of the Bland-Altman plots showed mean difference from HPE readings with a distribution of a large number of points at or near zero conforming with post NAST treated imaging measurements.

**Table 3:**
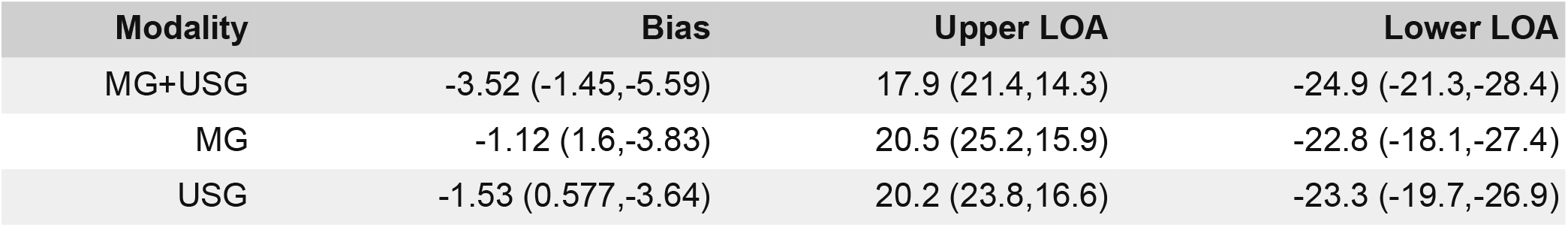
Bland Altman analysis of MG+USG Modalities: Bias and Upper and Lower limits of Agreement with 95% confidence limits.

| Modality | Bias | Upper LOA | Lower LOA |
| --- | --- | --- | --- |
| MG+USG | -3.52 (-1.45,-5.59) | 17.9 (21.4,14.3) | -24.9 (-21.3,-28.4) |
| MG | -1.12 (1.6,-3.83) | 20.5 (25.2,15.9) | -22.8 (-18.1,-27.4) |
| USG | -1.53 (0.577,-3.64) | 20.2 (23.8,16.6) | -23.3 (-19.7,-26.9) |

Despite the differences in radiological and HPE measured tumour sizes, T status remained unchanged in 71/109 (65.1%) cases. Smaller tumours (ypT0 and ypT1) appeared to have lower concordance compared to larger tumours. Maximum concordance was noted for ypT3 status (87.5%) followed by ypT2 status (77.8%), ypT0 status (58.8%) and ypT1 status (51.6%). However, there were only 8/109(7.3%) tumours with ypT3 status compared to the number of tumours with ypT0-ypT2 status **(Table 2 and Figure 2c)**. Concordance rate of ypT0 tumours was significantly different from rest of the classes **(Supplementary Table)**.

34 out of the 109 cases (31.2%) had no residual invasive tumour on final HPE (ypT0), one of which showed residual DCIS (ypTis) while 31(28.4%) patients had pCR. Out of the total cases with pCR, 18(58.1%) cases were correctly predicted by USG. While, 71/78 (91.0%) cases in which residual disease was noted on final HPE (no pCR), were correctly predicted as residual disease by USG **(Figure 3a) (Table 2)**. Thus, USG had sensitivity 91%, specificity 58.1%, PPV 84.5% and NPV 72% (**(Table 4)**.

**Table 4:** Sensitivity Analysis of various features of residual tumour by different imaging modalities.

| Feature | Modality | Sensitivity (%) | Specificity (%) | PPV (%) | NPV (%) | Unweighted Kappa | Accuracy (95%CI) | N |
| --- | --- | --- | --- | --- | --- | --- | --- | --- |
| Residual Disease | USG | 91 | 58.1 | 84.5 | 72 | 0.521 | 0.72<br>(0.731-0.884) | 109 |
| Residual Disease in Hormone positive | USG | 95.1 | 83.3 | 97.5 | 71.4 | 0.732 | 0.714<br>(0.825-0.987) | 47 |
| Residual Disease in HER2 enriched | USG | 83.3 | 53.8 | 62.5 | 77.8 | 0.367 | 0.778<br>(0.465-0.851) | 25 |
| Residual Disease in TNBC | USG | 88 | 50 | 78.6 | 66.7 | 0.406 | 0.667<br>(0.588-0.882) | 37 |
| Positive Node | USG | 64.6 | 91.8 | 86.1 | 76.7 | 0.579 | 0.767<br>(0.711-0.869) | 109 |
| Residual Tumor | MG+USG | 89.3 | 58.8 | 82.7 | 71.4 | 0.506 | 0.714<br>(0.711-0.869) | 109 |
| Residual Tumor | MG | 81.8 | 95.5 | 97.3 | 72.4 | 0.716 | 0.724<br>(0.757-0.936) | 66 |
| Residual Tumor | USG | 88.0 | 58.8 | 82.5 | 69.0 | 0.488 | 0.69<br>(0.7-0.861) | 109 |

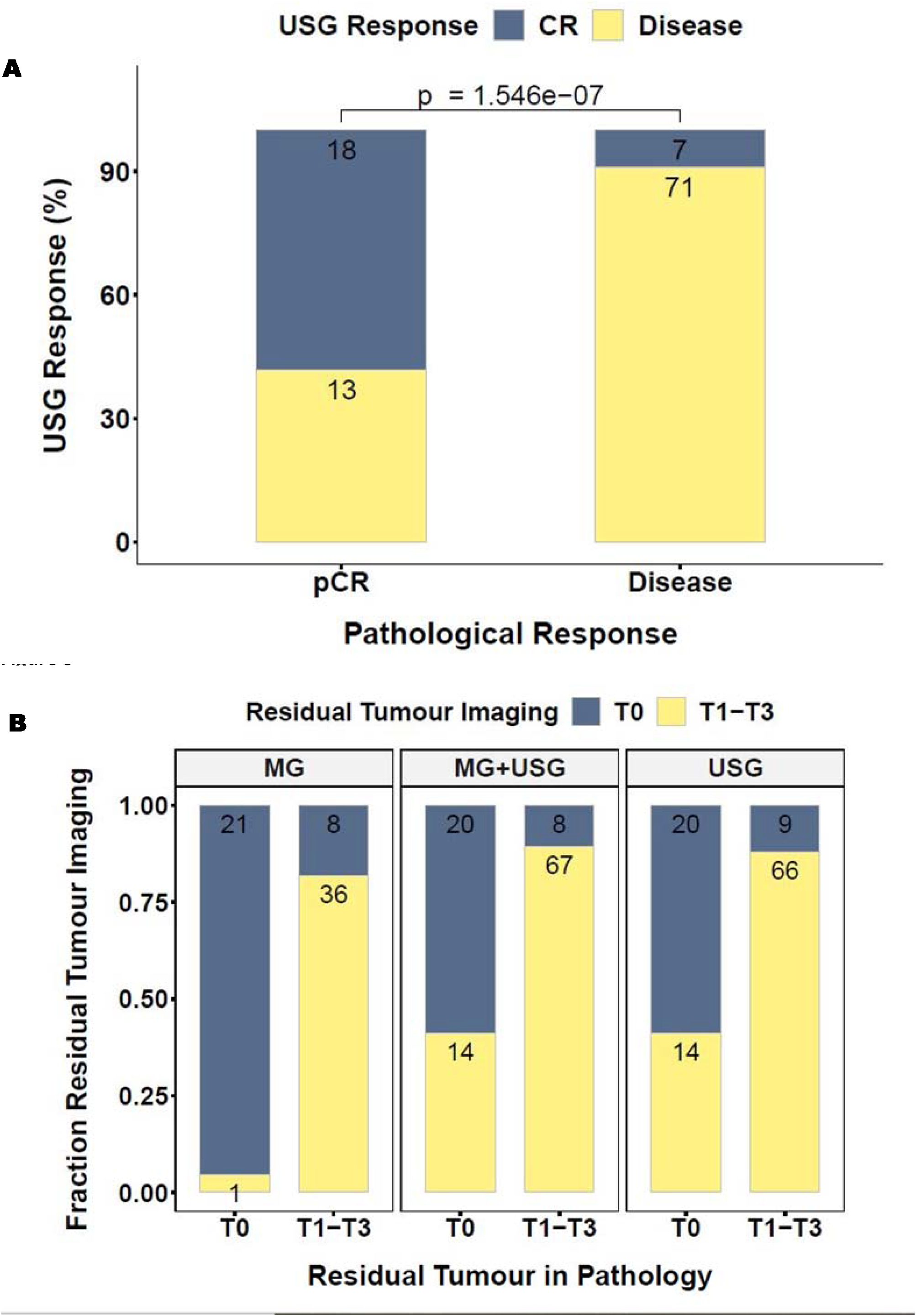

Sizes of the ypT0 tumours (pCR) that were overestimated by USG as residual disease in 14 cases, ranged between 6-37 mm (mean= 16.3 mm) and measured 0.1-10 mm for 4 patients, 11-20 mm for 7 patients, 21-30 mm for 1 patient and 31-40 mm for 2 patients.

MG had better ability to predict ypT0 with PPV 95.5% compared to USG (58.8%) but had slightly lower ability to predict residual disease with NPV 81.8% compared to USG (88%).[1] However, we found that USG had higher accuracy at 86.4% compared to 78.9% from MG. Therefore USG and MG might differ in prediction of residual disease (T0) but it was not possible to delineate with sample size n =66 for MG and 109 for USG. **(Figure 3B and Table 4)**.

Out of the total 61/109 cases (56%) with complete nodal response to NAST on final HPE (ypN0), 56 (91.8%) cases were correctly predicted by USG. 31/ 48 cases (64.6%) with axillary nodal metastasis were correctly identified on USG **(Figure 3c) (Table 2)**. Thus, USG had a sensitivity of 64.6%, specificity of 91.8%, PPV 86.1%, NPV 76.7% for predicting nodal metastasis with an unweighted kappa of 0.579 suggestive of fair to good agreement **(Table 4)**.

Radiology-pathology concordance for longest tumour dimension was noted in 30 out of 47 (63.8%) HR positive tumours, 21 out of 37 (56.8%) TNBCs and only 12 out of 25 (48.0%) HER2 enriched tumours **(Figure 4a and Table 2)**. Only 6/47 HR positive tumours (12.8%) showed pCR while 12/37 TNBCs (32.4%) and 13/25 HER2 enriched tumours (52.0%) showed pCR (Case 1). **Table 4 and figure 4b** depict the performance of USG in predicting residual disease/pCR among these molecular subtypes. However, the differences between the subtypes for distribution of concordance of longest dimension and prediction of pCR were not statistically significant (p value 0.43 and 0.46 respectively).

**Figure 4.**
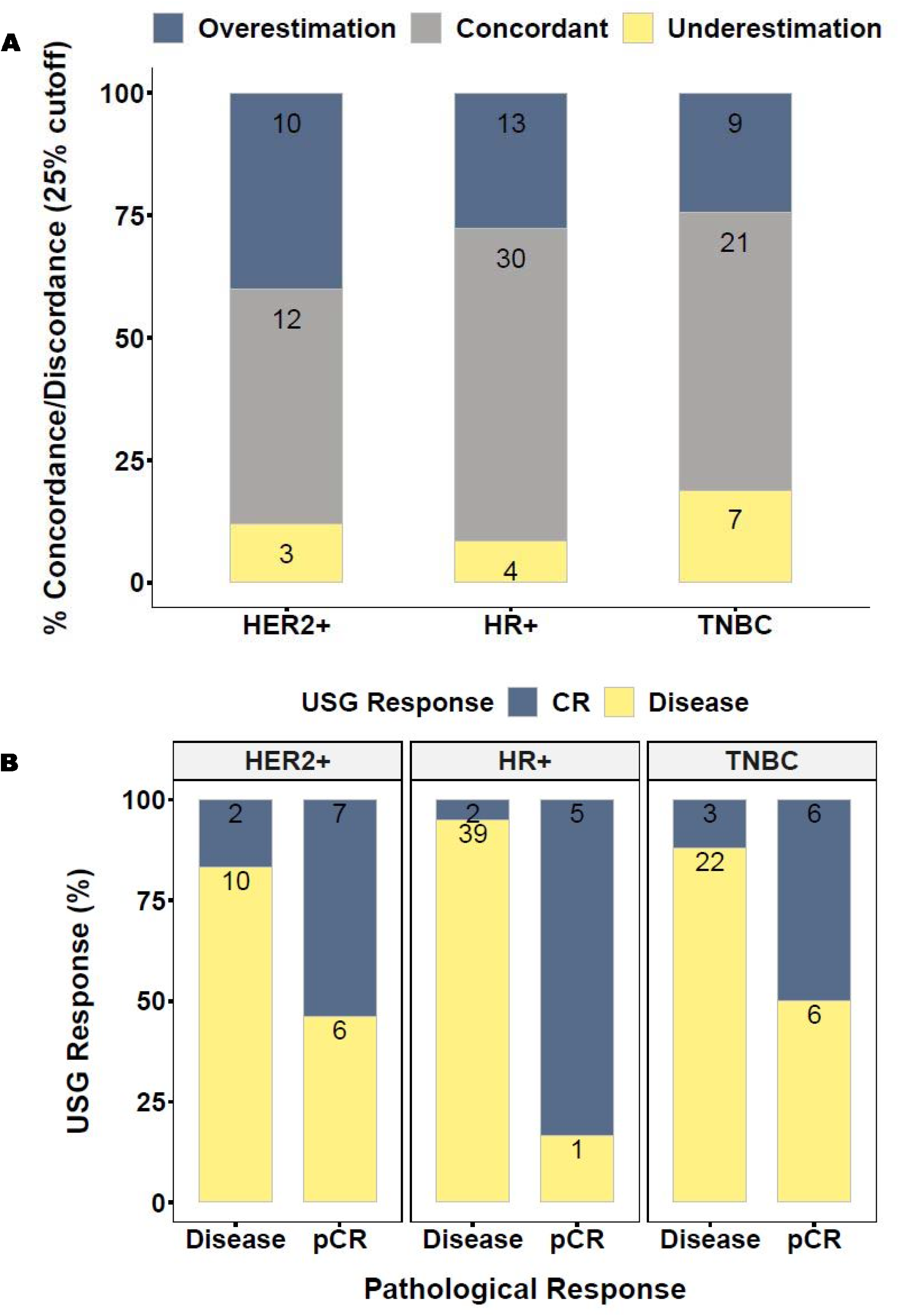

In our study, out of 57 cases without any residual DCIS on final HPE, 12(21.1%) cases showed persistent calcifications on MGs (**Case study 4)**. On the other hand, out of 13 cases with DCIS on final HPE, 7(53.8%) cases did not show calcifications on MG. In most of these cases DCIS was noted in the form of small foci ranging from 1-5 mm in extent, while only one case showed significant DCIS measuring 16 mm in extent (**Table 2) [supplementary figure-2/DCIS-Mammography]**.

Amongst the cases with discordance between imaging HPE tumour sizes, T status was unchanged in 15/ 46 discordant (32.6%) cases. **(Summary in Table 5)**. Out of the 46 discordant cases, 24 patients underwent large excision volume (LEV) surgeries. Out of these, 12 were volume replacement (VR) surgeries with >50% of breast volume resection, 6 being partial mastectomies followed by immediate implant reconstruction/VR with implant and 6 were VR with local/regional flap reconstructions like latissimus dorsi flap (LDF) or perforator flap (PF). Remaining 12 were OBS with 20-50% of the breast volume resection/volume displacement (VD) level 2. Out of these, in 12 cases discordance between imaging and pathology could have led to LEV surgeries as the imaging did not predict the tumour size accurately. Remaining 22 patients underwent either BCS (8 cases) or simple OBS with < 20% of the breast volume resection (VD Level 1) (14 cases).

**Table 5:** Large Excision Volume Surgery in Discordant cases (n=24)

| Type of Surgery | Reason for LEV surgery | Imaging size(mm) | Final HPE size (mm) | LEV surgery due to discordance | No<br>F<br>(n=24) |
| --- | --- | --- | --- | --- | --- |
| VR-Implant | Discordance, small breast, skin involvement | 23 | 10 | Yes |  |
| VR-Implant | Extensive residual microcalcifications | 22(ycT),100 (cal) | 15(ypT),90 (Tis) | No |  |
| VR-Implant | Extensive residual microcalcifications | 0(ycT),80 (cal) | 15(ypT), No Tis | No |  |
| VR-Implant | Patient choice | 16 | 2 | No |  |
| VR-Implant | Large tumour, no response to NACT | 80 | 51 | No* |  |
| VR-Implant | Small breasts, TNBC, RA, contraindications to RT, Contralateral breast DCIS | 18 | 30 | No |  |
| VR-LDF | Discordance | 45 | 32 | Yes |  |
| VR-LDF | Discordance | 34 | 22 | Yes |  |
| VR-LDF | Discordance | 37 | 0 | Yes |  |
| VR-LDF | Discordance, small breast, skin involvement, incomplete response to NACT | 36 | 21 | Yes |  |
| VR-PF | Cosmesis | 0 | 10 | No |  |
| VR-PF | Cosmesis | 0 | 5 | No |  |
| VD-Level 2 | Extensive residual microcalcifications | 56(ycT),40 (cal) | 25(ypT),2.5 (Tis) | No** |  |
| VD-Level 2 | Extensive residual microcalcifications | 12(ycT),45 (cal) | 0(ypT),2 (Tis) | No |  |
| VD-Level 2 | Extensive residual microcalcifications | 0(ycT),40 (cal) | 10 (ypT),0(Tis) | No |  |
| VD-Level 2 | Discordance | 25 | 8 | Yes |  |
| VD-Level 2 | Discordance | 30.7 | 1 | Yes |  |
| VD-Level 2 | Discordance | 43 | 0 | Yes |  |
| VD-Level 2 | Discordance | 17.7 | 0 | Yes |  |
| VD-Level 2 | Discordance | 13.3 | 28 | Yes |  |
| VD-Level 2 | Discordance | 17.2 | 8 | Yes |  |
| VD-Level 2 | Discordance | 16.5 | 8 | Yes |  |
| VD-Level 2 | For cosmetic purpose due to large ptotic breasts | 6 | 0 | No |  |
| VD-Level 2 | For cosmetic purpose due to large ptotic breasts | 0 | 3 | No |  |

\* decision of LEV surgery was not affected by discordance
\*\* decision of LEV surgery was due to extensive residual microcalcifications, not due to discordance in residual tumour size

Analysis of 9 cases with a wider discordance (maximum difference of 43 mm) of residual imaging size and final HPE size showed, 3 cases had fibrosis on final HPE without any residual lesion, 1 case showed fragmented response to NACT, 2 cases had residual calcifications on imaging, while 2 patients had skin involvement. In one case, the tumour size was accurately predicted by USG while it was overestimated by MG due to indistinct margins of the tumour and peritumoral architectural distortion, making estimation of the tumour size difficult.

Out of 109 cases, 8 cases (7.3%) had positive margins after excision on the initial frozen section which were revised intraoperatively. All these revised margins were assessed again by frozen sections immediately and were found to be negative for all of these cases. We also noted that none of the cases with positive margins on the initial frozen section had radiology-pathology discordance.

After a median follow-up of 19.1 months, there were 14 recurrences (10 distant, 2 local and 2 locoregional) and 5 deaths. In the overall cohort, recurrence free survival (RFS) and overall survival (OS) rates at median follow up were 87.2% and 95.4% respectively **(Table 1)**. Out of total 14 recurrences, 7 cases (50%) had discordant imaging. 8 of these 14 patients with recurrences (57.1%) had TNBCs. None of these cases had positive margins after excision on the frozen section. Out of 5 deaths, only 1 case with TNBC tumour (20%) had local recurrence which had radiology-pathology concordance, 3 cases (60%) had distant metastases while in 1 case (20%) death was due to non-oncological comorbidities.

## DISCUSSION

Several studies have compared performance of physical examination and diagnostic imaging modalities like MG, USG and MRI for assessing tumour response to NAST. Some studies have demonstrated better performance of USG compared to MG (Leddy *et al*. 2016; Cortadellas *et al*. 2017; Yang *et al*. 2020) while studies by Gruber et al, Ramirez et al have demonstrated better performance of MG compared to USG (Ramirez *et al*. 2012; Gruber *et al*. 2013). On the other hand, Herrada et al has reported that predicting pCR is not highly accurate with either MG or USG (Herrada *et al*. 1997).

Several studies have shown that MRI showed a better correlation with the pCR and pathological residual size after NAST than CBE, MG or USG (Warren *et al*. 2004a; Shin *et al*. 2018; Partridge *et al*. 2018). In a study performed by Scheel et al (ACRIN6657), although the longest diameter measurements of residual disease by MRI more closely matched final HPE size than did those by MG or by CBE, the correlation was low (r = 0.33 for all lesions) (Scheel *et al*. 2018). However, it was within the wide range of those previously reported (range, 0.21–0.89) (Rosen *et al*. 2003; Bhattacharyya *et al*. 2008; Chen *et al*. 2011). MRI is known to underestimate residual disease as anti-angiogenic drugs used during NAST can inhibit contrast uptake during dynamic contrast enhanced MRI studies. Also post NAST tumor fibrosis cannot be reliably differentiated from residual tumour and can lead to overestimation of tumor sizes on MRI (Rosen *et al*. 2003; Warren *et al*. 2004b; Yeh *et al*. 2005; Naglaa Abdel Razek 2013). Chen et al has reported that the limitation of MR imaging in detecting the minimal residual disease may impact surgical management (Chen *et al*. 2011). Although MRI technology is evolving over time, relatively lower accuracy of MRI was observed even in the more recent studies also including a meta-analysis of preoperative MRI (Houssami *et al*. 2008). The high relative cost of MRI, combined with potential advantages of clinical examination and ultrasound in terms of accessibility, suggest that a combination of the latter may be a reasonable alternative testing strategy to MRI in preoperative assessment after NAST (Marinovich *et al*. 2013). Besides MRI has to be performed at least twice during the NAST regimen which increases the economic burden on the patient which could otherwise be reserved for the purpose of treatment.

The concordance rate of MG (68.2%) was better than USG (52.3%) as we observed (**Figure 2a and Table 2)** is correlated with the concordance rates for MG and USG considering 25% as cut-off were 60.5% and 54.3% respectively, reported by (Lai *et al*. 2016). However, in our study, the difference was not statistically significant (p value 0.081). On the other hand, in the study by Ramirez et al, MG measurements most closely correlated with pathological measurements compared to USG and MRI (Ramirez *et al*. 2012). Gruber et al did not find any significant difference between MG and HPE sizing while tumour size was significantly underestimated by the USG (Gruber *et al*. 2013). Combination of both the modalities had a concordance rate of 57.8% in our study while it was comparatively better (69.1%) in the study by Peintinger et al. Comparisons of MG and USG in the same patients showed similar concordance rates, suggesting comparable performance between the two modalities (Peintinger *et al*. 2006). Smaller tumours (ypT0 and ypT1) appeared to have lower concordance compared to larger tumours **(Table 2 and Figure 2c)**. This was contrary to the higher concordance seen with smaller tumours in the study by Hamza et al (Hamza *et al*. 2018). which can be attributed to the residual post NAST fibrosis and fragmentation seen in these tumors in our study. In cases showing fibrosis on final HPE **(Case study 3)**, we observed that the imaging interpretation was difficult as there was a reduction in the density of these lesions on the MG and a more iso to hypoechoic appearance of these lesions on the USG with an ill-defined dendritic appearance. Moreover, in 8 cases in which the imaging showed complete resolution of the tumour (ycT0), final HPE revealed ypT1 tumours, the difference in size being in the range of 3-15 mm which did not significantly impact the surgical decision.

In our study, USG showed good ability for prediction of residual disease with PPV of 84.5% which correlated well with the study by Keune et al (85.2%) (Keune *et al*. 2010) and data synthesis of 6 studies done by Croshaw et al (85%) (Croshaw *et al*. 2011). NPV for pCR prediction (72%) also resembled with Keune et al (68.8%) (Keune *et al*. 2010) and was better than data synthesis by Croshaw et al (44%) (Croshaw *et al*. 2011). The accuracy of USG was 72%, lower than that found by Peintinger et al (88.9%) (Peintinger *et al*. 2006). An unweighted Kappa of 0.521 suggested fair to good agreement between imaging and pathologic data similar to Keune et al (0.50) (Keune *et al*. 2010) **(Table 4)**. The avoidance of surgery remains only a future goal for patients in whom an absence of residual tumor can be accurately detected. While imaging advances aim at improving the accuracy of pCR prediction, which is a predictor(biomarker) for improved disease-free survival and overall survival, it is less likely to benefit from an elimination of surgery after NAST.

In our study, the tumors that were underestimated as complete response by USG and MG measured 0.3-1.5 cm at pathology, while in meta-analysis by Marinowich et al (Marinovich *et al*. 2015) using MRI, the tumours measured 0.1-11cm. In our study, USG measurements of cases with pCR ranged between no residual tumour (18/31, 58.1%) and 0.6-4.3 cm (mean= 2.1±1.8 cm) and size of > 2 cm occurred in 3/31(9.7%) cases. While in meta-analysis 2015 by Marinowich et al (Marinovich *et al*. 2015), the range was 0.3-6.1 cm (median-2 cm) and the size of > 2 cm was noted in 8/57(14%) cases which is not statistically significantly different from our study (*p*=0.74, CI=95%, Fisher’s exact test for count data).

We found that USG performed well in predicting nodal metastasis PPV of 86.1% and NPV of 76.7%. One of the notable advantages of ultrasound is its ability to evaluate axillary nodes. A pre-NAST documentation of > 2 abnormal axillary nodes guide the surgeon on the necessity of an ALND and avoids the unnecessary procedure of a SLNB. The possibility of clinically underestimating disease burden in this scenario is also avoided. Even if there are no palpable nodes in the axilla, USG of the axilla helps to identify patients with abnormal appearing nodes. USG guided biopsy of an abnormal appearing node can then be done to confirm the presence of metastatic disease. Such patients may then be triaged for NAST. SLNB is reserved for patients with T0-T2 tumours showing complete nodal responses or with 2 or less abnormal nodes on imaging after NAST.

In our study, the least concordance rate was noted in HER 2 enriched tumours (48%) compared to HR positive tumours (63.8%) and TNBCs (56.8%). Similarly, although the maximum percentage of cases showing pCR were seen in HER2 positive tumours due to targeted anti-HER2 therapy, imaging prediction of residual tumour was less accurate (PPV of 62.5%) for these types of cancers, though NPV was better 77.8% **(Table 4)**. Retrospective analysis of these cases showed higher incidence of post-treatment fibrosis in these cases which was overestimated as residual tumour on imaging **(Case study 3)**. Indistinct margins and associated microcalcifications were also more commonly observed in these cases leading to difficulties in estimating exact tumour size. In the study conducted by Yoo et al statistically significant worse correlation was found between the tumor size measured by imaging and pathology in HER2 positive molecular subtype compared with luminal A and luminal B subtypes (p¼0.008 and 0.007, respectively) (Yoo *et al*. 2017). On the other hand, in our study, despite the differences in the distribution of the concordance for longest dimension as well as prediction of pCR among different histologic subtypes, these differences were not statistically significant (p value 0.43 and 0.46 respectively). These findings were similar to the findings noted by Scheel et al (ACRIN6657) which did not identify any influence of histologic subtype on the associations between preoperative measurements and pathology size or the accuracy for detecting pCR (p = 0.14–0.83) (Scheel *et al*. 2018). On the other hand, the study conducted by Waldrep et al noted that the accuracy of predicting presence or absence of pCR by imaging varies by breast cancer subtype (Waldrep *et al*. 2016).

Out of the cases without residual DCIS on final HPE, 21.1% cases showed persistent calcifications on MG **(Case study 4)**. Presence of microcalcifications on post-NAST imaging is therefore not a reliable indicator of persistence of residual tumour after NAST as was observed in our study. Microcalcifications of a ductal carcinoma may persist in cases of good response and calcifications secondary to necrosis may appear (Ollivier *et al*. 2005; Li *et al*. 2014). The cases with residual DCIS on final HPE, not presenting as calcifications on MG, only one case showed significant DCIS measuring 16 mm in extent which could impact the surgical decision. It is known that non-calcified DCIS may not be seen on conventional imaging and requires investigation like MRI for detection as shown in studies by (Gruber *et al*. 2013; Kuhl *et al*. 2017).

Retrospective analysis of the surgical outcomes revealed that in 12 out of 46 discordant cases (26%), the discordance between imaging and pathology could have led to LEV surgeries as the imaging did not predict the tumour size accurately **(Table 5)**. The question this raises is that does this radiological discordance adversely affect the surgical decision? Assuming that surgeons consider the imaging-determined measurement when planning resection, such overestimation would lead to unnecessarily large excision. Nevertheless, imaging appearances cannot be ignored and form an important part of the surgical decision. Gampenrieder et al has stated that removal of the radiological residual area of the primary tumor is considered standard care after NACT as it represents the tumour bed (Gampenrieder *et al*. 2019). Therefore, imaging findings provide guidance to the surgeon regarding the surgical approach and the extent of the excision. It should also be noted that the surgical decision depends on a number of factors, the disease burden at presentation, patient’s breast volume, the response of the disease to NAST, pre and post NAST imaging features, presence of DCIS, molecular subtype, patient and surgeon preferences and also the desired cosmesis. Even small tumours in patients with small breast volumes may need excision followed by volume replacement procedure like PF reconstruction. On the other hand, patients with large breast volumes may be managed with BCS or simple OBS.

In our study, out of 8 cases that had positive margins after excision on the initial frozen section, all the cases showed negative revised margins on immediate intraoperative frozen section. The absence of re-operation cases noted in our study could be attributed to the use of intraoperative frozen section. On the other hand, the re-operation rate found in the study by Bhattacharya et al due to positive margins was 9.5% which had used MRI for preoperative imaging (Bhattacharyya *et al*. 2008). One peculiar observation we noted was that none of the cases with positive margins on the initial frozen section had radiology-pathology discordance. We could not find any explanation for the same.

In the overall cohort, RFS and OS rates at median follow up of 19.1 months were 87.2% and 95.4% respectively. In the study by Gampenrieder et al using MRI for preoperative imaging, RFS and OS rates were 82.9% and 89.4% respectively after median follow up of 31 months (Gampenrieder *et al*. 2019).

Limitations of the study are relatively smaller sample size and retrospective design of the study. MG was not available in 39 patients. MRI was reserved only for specific cases. We did not collect data on markers of proliferation, such as Ki-67, that would allow further characterization of HR positive tumors into luminal A and B subtypes and facilitate analyses of differences between these subsets in imaging accuracy. The median follow-up period was quite short. This resulted in a relatively small number of recurrences, making definitive conclusions about survival outcomes difficult. As apart from imaging features, the surgical decision depends also on a number of other factors like patient and surgeon preferences which are difficult to assess, evaluation of the surgical outcomes of radiology-pathology discordance may not be accurate.

## FUTURE DIRECTIONS

Since studies have shown that contrast-enhanced mammography has comparable performance to contrast enhanced MRI for lesion detection and estimation of tumour size (Fallenberg *et al*. 2014, 2017; Lobbes and Houben 2016), recently after upgrading to contrast-enhanced mammography equipment, we have started its use in cases undergoing NAST for better assessment of residual tumour size and pCR and also as a more economical alternative to MRI. We also intend to perform elastography during all pre-NACT as well as post-NAST USGs.

## CONCLUSION

Our study highlights the use of cheaper and widely available conventional imaging modalities for predicting the pathological residual tumour size after NAST which is the need in the developing countries where the cost of treatment is to be contained. Both MG and USG showed comparable performance with moderate concordance with the pathology. USG showed excellent ability for prediction of the residual disease and complete axillary nodal response but overall lower ability to predict pCR and nodal metastasis. Although in our study, molecular subtypes were not found to influence the radiology-pathology concordance significantly, we noted that imaging prediction of residual tumour was less accurate for HER 2 enriched cancers due to higher incidence of post-treatment fibrosis in these cases. Also, calcifications seen on post NAST imaging may not correspond to residual disease. However, in these cases the imaging appearances were still used to guide the surgical plan. The surgical outcomes were fairly well managed in the discordant cases with the oncoplastic surgical techniques as these allow to excise larger volumes of breast tissue with tumour-free margins, fewer re-operations and mastectomies and still achieved good cosmesis with acceptable OS and RFS.

## Data Availability

The data available related to the manuscript will be available on request after consultation with the institutional ethical committee.

---

**Suppl Fig 1.**
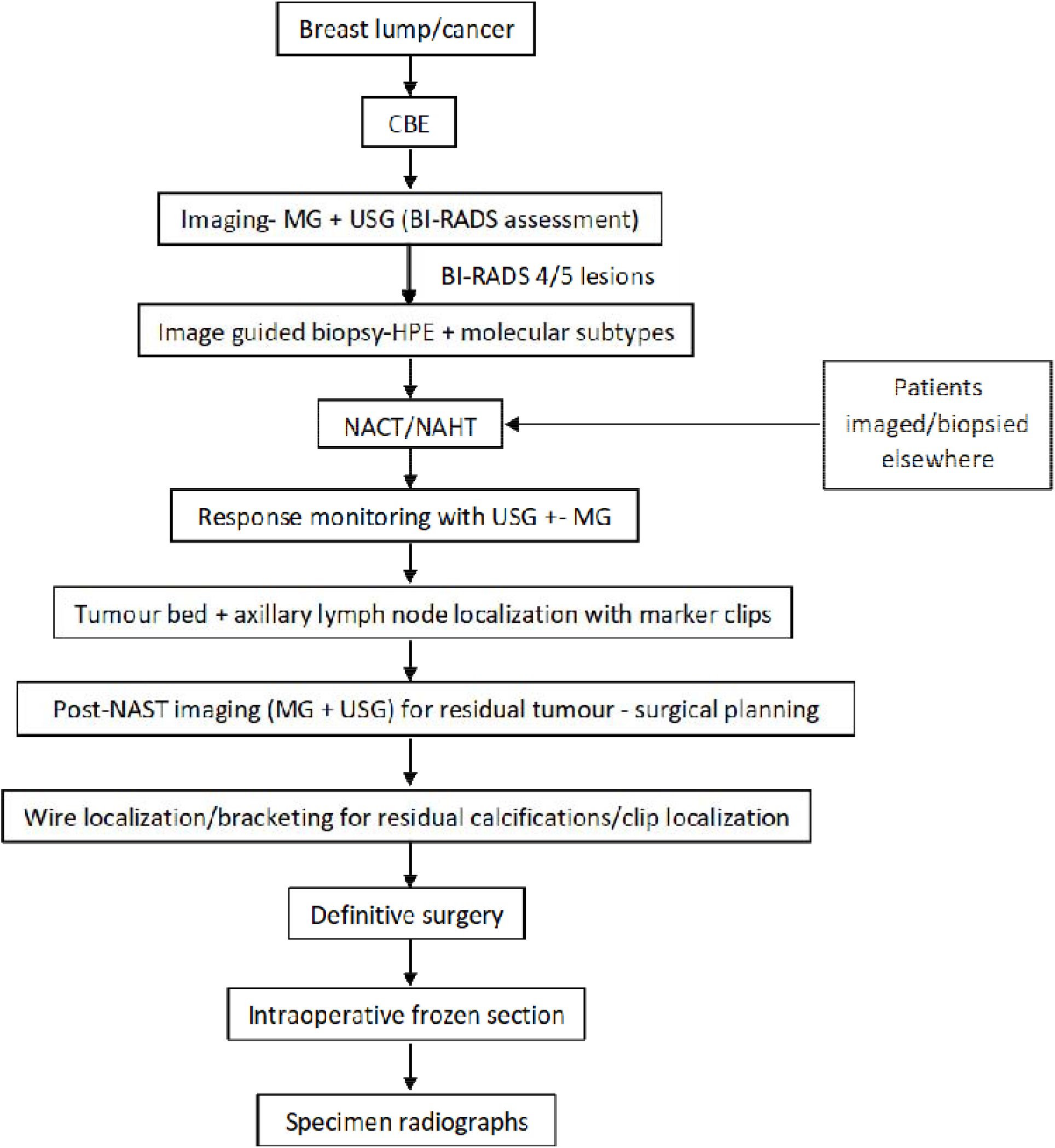

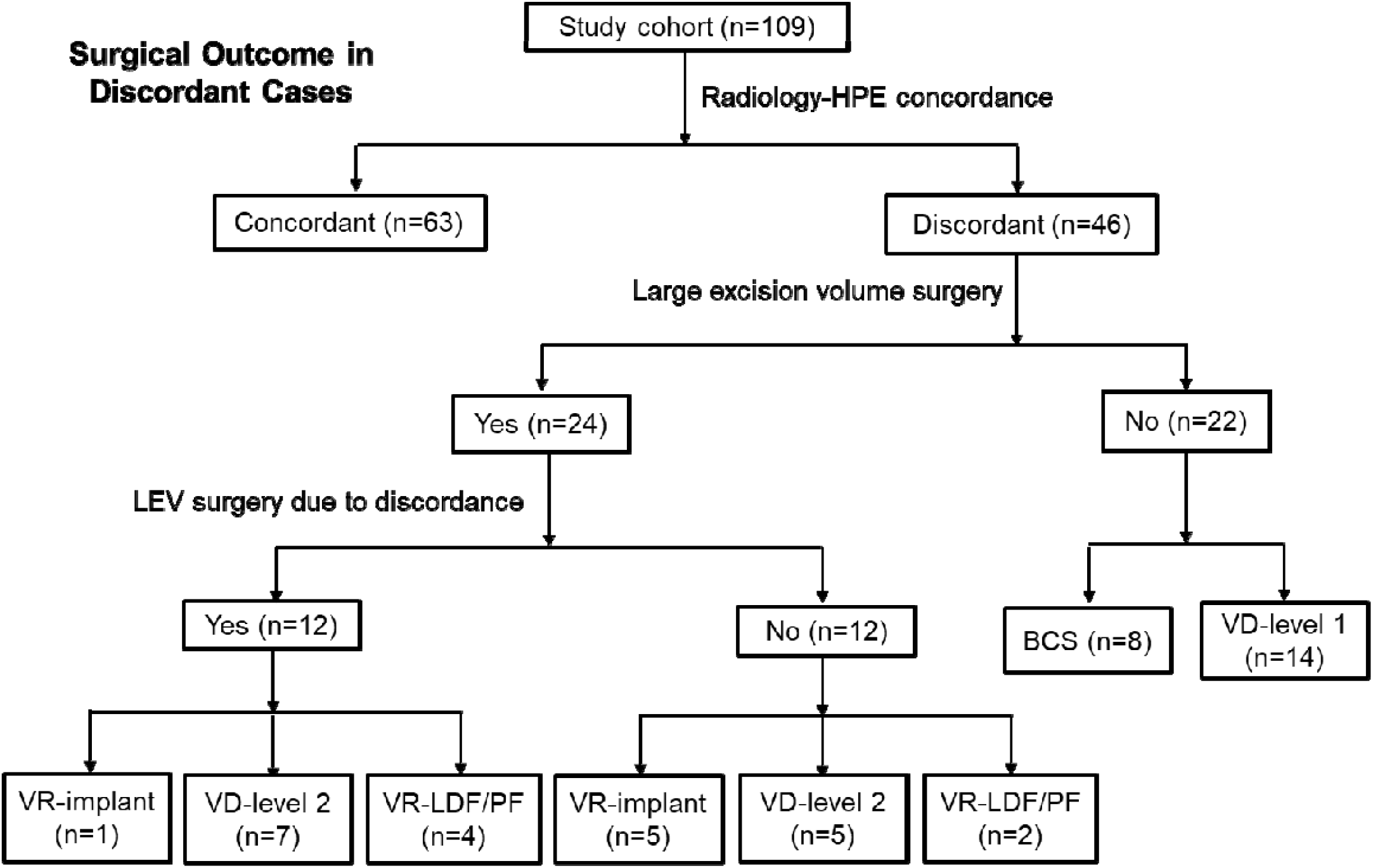

